# Modeling the COVID-19 Vaccination Dynamics in the United States: An Estimation of Coverage Velocity and Carrying Capacity Based on Socio-demographic Vulnerability Indices in California

**DOI:** 10.1101/2021.07.03.21259881

**Authors:** Alexander Bruckhaus, Aidin Abedi, Sana Salehi, Trevor A. Pickering, Yujia Zhang, Aubrey Martinez, Matthew Lai, Rachael Garner, Dominique Duncan

## Abstract

**Background:** Coronavirus disease 2019 (COVID-19) disparities among vulnerable populations are of paramount concern that extend to vaccine administration. With recent uptick in infection rates, dominance of the delta variant, and proposal of a third booster shot, understanding the population-level vaccine coverage dynamics and underlying sociodemographic factors is critical for achieving equity in public health outcomes. This study aimed to characterize the scope of vaccine inequity in California counties through modeling the trends of vaccination using the Social Vulnerability Index (SVI).

**Methods:** Overall SVI, its four themes, and 9228 data points of daily vaccination numbers from December 15, 2020, to May 23, 2021, across all 58 California counties were used to model the growth velocity and anticipated maximum proportion of population vaccinated, defined as having received at least one dose of vaccine.

**Results:** Based on the overall SVI, the vaccination coverage velocity was lower in counties in the high vulnerability category (v=0.0346, 95% CI: 0.0334, 0.0358) compared to moderate (v=0.0396, 95% CI: 0.0385, 0.0408) and low (v=0.0414, 95% CI: 0.0403, 0.0425) vulnerability categories. SVI Theme 3 (minority status and language) yielded the largest disparity in coverage velocity between low and high-vulnerable counties (v=0.0423 versus v=0.035, *P*<0.001). Based on the current trajectory, while counties in low-vulnerability category of overall SVI are estimated to achieve a higher proportion of vaccinated individuals, our models yielded a higher asymptotic maximum for highly vulnerable counties of Theme 3 (K=0.544, 95% CI: 0.527, 0.561) compared to low-vulnerability counterparts (K=0.441, 95% CI: 0.432, 0.450). The largest disparity in asymptotic proportion vaccinated between the low and high-vulnerability categories was observed in Theme 2 describing the household composition and disability (K=0.602, 95% CI: 0.592, 0.612; versus K=0.425, 95% CI: 0.413, 0.436). Overall, the large initial disparities in vaccination rates by SVI status attenuated over time, particularly based on Theme 3 status which yielded a large decrease in cumulative vaccination rate ratio of low to high-vulnerability categories from 1.42 to 0.95 (P=0.002).

**Conclusions:** This study provides insight into the problem of COVID-19 vaccine disparity across California which can help promote equity during the current pandemic and guide the allocation of future vaccines such as COVID-19 booster shots.

**Key Messages:**

- The Social Vulnerability Index (SVI) and its four themes along with the daily proportion of vaccinated individuals across the 58 California counties were used to model, overall and by theme, the growth velocity and anticipated maximum proportion of population vaccinated.
- Overall, high vulnerability counties in California had a lower vaccine coverage velocity compared to low and moderate vulnerability counties.
- The largest disparity in coverage velocity between low and high vulnerability counties was observed based on the SVI Theme 3 status (minority status & language).
- Based on the current trajectory, highly vulnerable counties based on SVI Theme 3 are expected to eventually achieve a higher proportion of vaccinated individuals compared to low vulnerable counterparts.
- Understanding the vaccine coverage dynamics and underlying sociodemographic factors is critical for achieving equity in public health outcomes during disease outbreaks and catastrophes.

## 1. Introduction

### Background

The coronavirus disease 2019 (COVID-19) has disproportionately impacted minority groups in terms of the number of cases and mortality rates, which further warrants equitable vaccine rollout to assist the communities in dire need of medical support^1^. Data from a Centers for Disease Control and Prevention (CDC) report covering county-level COVID-19 vaccination and social vulnerability revealed that in most states, vaccination coverage was higher in low and moderate vulnerability counties compared to high vulnerability counterparts^2^. A similar report across 40 United States (U.S.) states revealed that as of May 24, 2021, the percentage of White people who had received at least one dose of COVID-19 vaccine was about 1.5 times higher than Black people and 1.3 times higher than Hispanic people^3^. These data indicate widespread vaccination inequities across the U.S. that must be addressed to ensure that vulnerable populations receive vaccines at an equitable rate.

### Social Vulnerability Index (SVI)

Historically, population-level healthcare disparities have been attributed to the racial or ethnic composition of the community residents. However, with growing evidence becoming available, conceptual models of health inequity have evolved to include various other underlying dimensions such as sociodemographics, disability status, and geographic location^4^. Social Vulnerability Index (SVI), defined by the CDC, is a composite index composed of 15 census variables grouped into four main themes, resulting in 20 metrics each having a national and state-specific county ranking^5^. This metric was introduced to assist local authorities in the identification of vulnerable populations during disease outbreaks, natural disasters, or catastrophes. The four underlying themes include ‘socioeconomic status’, ‘household composition and disability’, ‘minority status and language’, and ‘housing type and transportation’^5^ (Figure 1).

**Figure 1.**
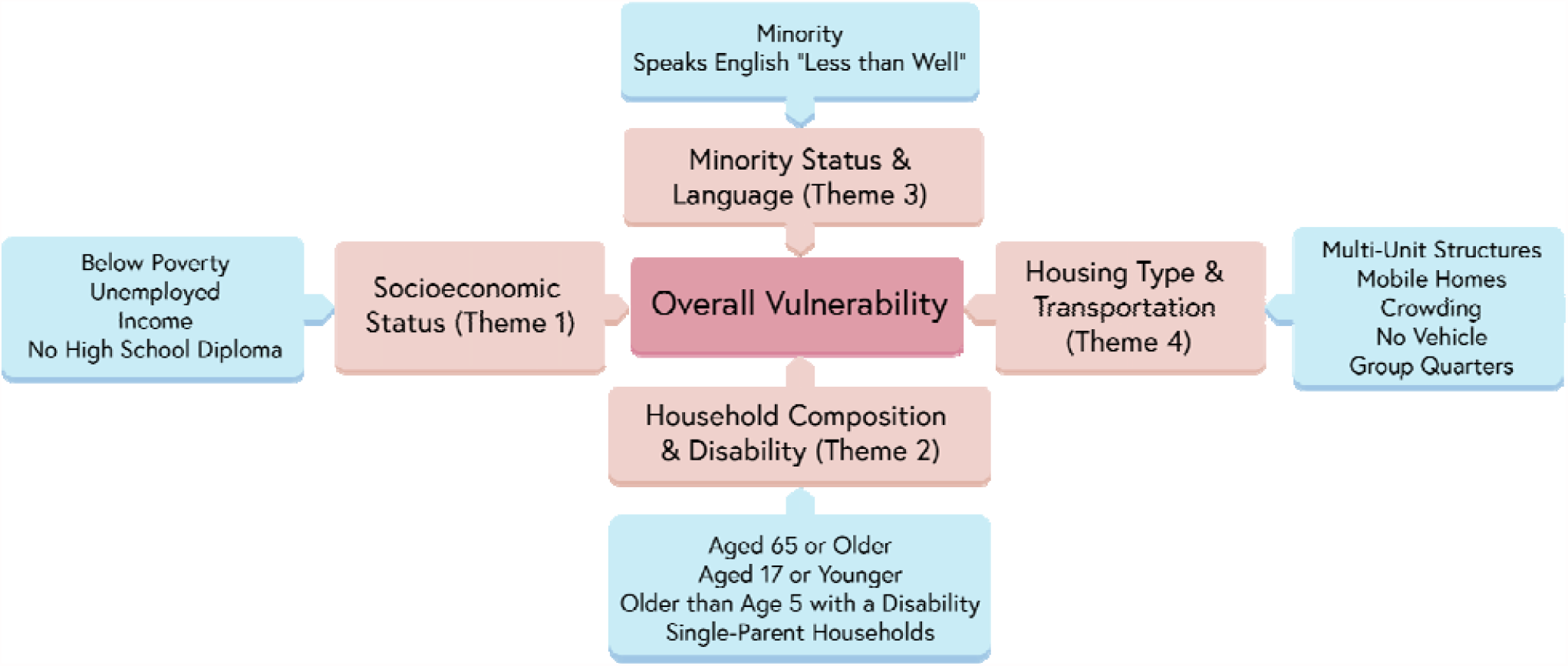
Social Vulnerability Index (SVI), underlying themes, and components

### Rationale and Objective

California, a highly heterogeneous state and the most populous in the U.S., has demonstrated the need for equity in vaccine rollout by setting aside 40% of doses for the hardest-hit communities^6^. Since this decision, an estimated 53% of individuals (as of May 4, 2021) in the lowest quartile of the Healthy Places Index remained unvaccinated as opposed to only 28% in the highest quartile^7^. This report further highlights the ongoing vaccine disparity problem in California. The major national and state-wide (California) vaccination events and policies are summarized in Figure 2. Although the gap in healthcare delivery among communities of different social vulnerability status is well-known, to the best of our knowledge, there is no peer-reviewed report on the dynamic trajectories of vaccination in relation to vulnerability indicators in the U.S. Therefore, in this study, we aimed to determine whether the longitudinal trends in vaccination rates across California counties differ by SVI. More specifically, we aimed to model the growth rate in vaccination coverage and the anticipated maximum proportion of vaccinated individuals in relation to county-level overall SVI and its underlying themes.

**Figure 2.**
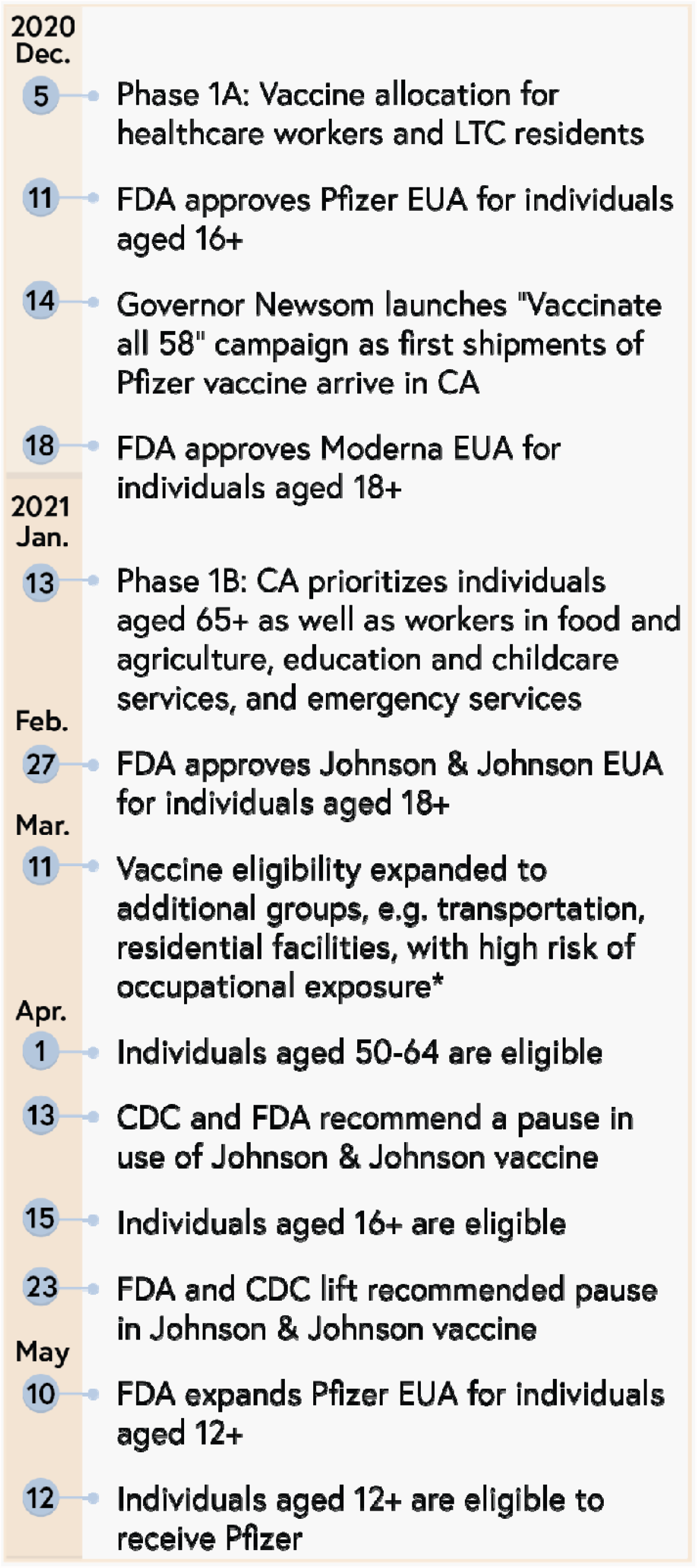
Chronological overview of major national and state-wide (California) vaccination events and policies. Prior to the arrival of the first shipment of Pfizer-BioNTech COVID-19 vaccines in California, the California Department of Public Health (CDPH) issued a statement on December 5, 2020, recommending an initial Phase 1a vaccine allocation for healthcare workers and older or at-risk residents of long-term care facilities. Vaccine supply was limited in the early stages of distribution, and these populations were determined to be at risk of direct exposure to the virus and were prioritized to receive the vaccine first^34^. The recommendation was made anticipating the federal government’s vaccine distribution efforts across the nation as the FDA issued Emergency Use Authorizations (EUA) for the Pfizer-BioNTech vaccine on December 11, 2020^35^ and for the Moderna COVID-19 vaccine on December 18, 2020^36^. At the time, these vaccines were approved for individuals aged 16 and older and for individuals aged 18 and older, respectively. With the first shipment of Pfizer-BioNTech vaccines, California Governor Gavin Newsom launched the “Vaccinate all 58” campaign on December 14, 2020 in an effort to distribute vaccines safely, fairly, and equitably for all 58 counties of the state^37^. On January 13, 2021, CDPH issued a statement opening up vaccine eligibility to individuals aged 65 and older. This population was identified as part of Phase 1b, and while vaccine demand was still much higher than the supply, the demand among healthcare workers declined among the initial Phase 1a populations^38^. Phase 1b populations included individuals aged 65 and older as well as workers in food and agriculture, education and childcare services, and emergency services^39^. Because the state gave counties the power to decide when to adopt vaccine phases based on vaccine availability, parts of California moved to Phase 1b earlier than others, and the various populations in Phase 1b were eligible on different dates across the state. A third vaccine, Johnson & Johnson/Janssen COVID-19 vaccine, was issued an EUA by the FDA on February 27, 2021 for individuals 18 and older^40^. This further increased vaccine availability in California as vaccine demand continued to outpace supply. A brief pause in the Johnson & Johnson/Janssen vaccine starting April 13, 2021^41^ was issued jointly by the FDA and CDC as a result of concerns that the vaccine led to six rare cases in the U.S. of blood clot development, but this pause was lifted on April 23, 2021 jointly by both agencies after a safety review determined that the potential benefits of receiving the Johnson & Johnson/Janssen vaccine outweighed the potential risks^42^. Further updates to vaccine eligibility for Californians were made in the next two months. On March 11, 2021, vaccine eligibility was extended to residents and workers in facilities such as prisons and homeless shelters (including the homeless population). These populations were determined to be in facilities at great risk of spreading the virus and around individuals likely to have medical conditions that increased the risk of developing negative effects from the virus. Eligibility was also extended to public transport, airport, and commercial airline workers because of their risk of contracting the virus at work. These workers were also determined to be working critical operations^43^. On April 1, 2021, individuals aged 50-64 were eligible to receive vaccines, and Californians aged 16 and older were eligible shortly after beginning April 15, 2021. The Moderna and Johnson & Johnson vaccines were still approved for individuals 18 and older only, but the FDA expanded the EUA on the Pfizer-BioNTech vaccine May 10, 2021 for vaccinating individuals aged 12 through 15^44^. This was followed by CDPH announcing vaccine eligibility for Californians aged 12 and older beginning May 12, 2021^39^. (LTC: Long-term care, FDA: U.S. Food and Drug Administration, EUA: Emergency Use Authorization, CDC: U.S. Centers for Disease Control and Prevention)

## 2. Materials and Methods

### Data Acquisition

The SVI values for California were obtained from the Agency for Toxic Substances and Disease Registry (ATSDR, 2018 data)^8^ and used to measure the levels of vulnerability across California’s 58 counties. These values differ from the nationwide data since the ranking is in reference to California counties. Each county is assigned a numerical value from 0-1 for the overall SVI level and its four themes^5^ (Figure 1). A value of 0 indicates the least vulnerability whereas a value of 1 indicates the highest vulnerability level. Counties were categorized into tertiles for overall SVI and each of its four components: SVI<0.33 describing low vulnerability, 0.33≤SVI<0.66 describing moderate vulnerability, and SVI≥0.66 describing high vulnerability. The vaccination data including the cumulative number of residents who received at least one vaccine dose was collected from the California Health & Human Services Agency (CHHS), which is a daily-updated dataset containing vaccination data of all 58 counties^9^.

### Data Curation

R version 4.0.4 and the tidyverse package were used to clean and curate the data. To produce the daily proportion of residents vaccinated in each county (p), the cumulative count of residents with at least one dose of any of the Moderna, Pfizer-BioNTech, or Johnson & Johnson/Janssen vaccines was used as the numerator. Similar to the CDC’s methodology^2^, the denominator was defined as the total population of each California county from the U.S Census Bureau’s 2019 estimates. For dates with missing vaccination data, we used the last available date’s value to impute the cumulative daily proportion of residents vaccinated for the respective county. Then, we merged each county’s respective SVI values, along with their SVI tertile ranks into the dataset. The final dataset contained one data point per county per day for all 58 California counties with a temporal coverage from December 15, 2020 (or the earliest available data) to May 23, 2021.

### Modeling Strategy and Statistical Analysis

Initial analyses were performed to determine the shape of the longitudinal curves of cumulative vaccination status (total with 1+ dose and total fully vaccinated) for each county. These curves followed that of a logistic growth curve, such as those traditionally used in ecology to describe the growth of a population towards an asymptotic value. This curve describes the value of the population at a particular time as a function of a per capita growth rate (“vaccination velocity”) and the carrying capacity (“asymptotic maximum”)^10^:

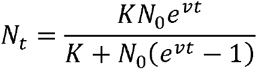

Using this framework to describe the number of individuals vaccinated, the instantaneous change in the population vaccinated at a given time can be described as:

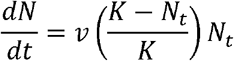

To fit these equations to our data, we used a nonlinear least-squares model fitting approach using the nls function in R^11,12^. This approach estimates the parameter values in the specified equation that best fit the data. Because we were interested in determining whether certain parameters (K and v) varied based on SVI status, we included dummy variable interactions in the model to allow for these parameter estimates to differ among SVI groups. To standardize the outcome across counties with different population sizes, we used the proportion of individuals vaccinated (p) as the outcome. Because all counties started with a very low proportion of individuals vaccinated, we treated the starting proportion p_0_ as a nuisance parameter and did not test for differential effects in p_0_ by SVI status. We constructed SVI indicator variables: i_1_ being an indicator of a county having “moderate” SVI status and i_2_ being an indicator of a county having “high” SVI status. With dk_1_ and dk_2_ capturing the difference in the k term for counties with Moderate and High SVI status, respectively, and dv_1_ and dv_2_ capturing the respective differences in the v term, the final equation we fit was:

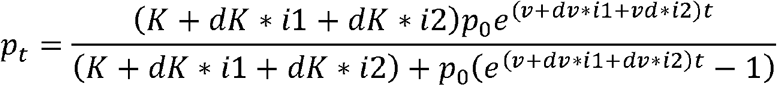

Estimated v and K parameters for low, moderate, and high SVI counties were computed using the lincom function in the biostat3 package.

To determine whether vaccination disparities by SVI status in early 2021 persisted through time, we used negative binomial regression models to examine the rate ratios of proportion vaccinated in low and moderate versus high SVI counties at three separate “snapshots” in time: January 1, March 1, and May 1, 2021. In these models we used cumulative number of individuals vaccinated as the outcome, with an offset of the natural log of county population size, thus effectively modeling the rate of individuals vaccinated at each county. Models included an interaction term between SVI category and date, to test whether the proportional differences in vaccination rates persisted throughout the study period.

## 3. Results

### Data Summary

A total of 9228 data points from 58 California counties were included in the analysis. There were 258 missing data points (2.79%), many of which fell on holidays and weekends when a lag in reporting likely occurred. The distribution of vaccination coverage at study endpoint and overall SVI ranks are depicted in Figure 3. The average county population estimates were comparable among the counties within the three categories of overall SVI (*P-*value range: 0.76-0.1, supplementary figure S1). At the study endpoint, the average proportion of vaccinated population in counties with low, moderate, and high overall SVI was 0.54±0.10, 0.48±0.13, and 0.40±0.08, respectively, with a lower proportion in high vulnerability category as compared to moderate (*P*=0.0351) and low (*P*=0.0003) categories (supplementary figure S1).

**Figure 3.**
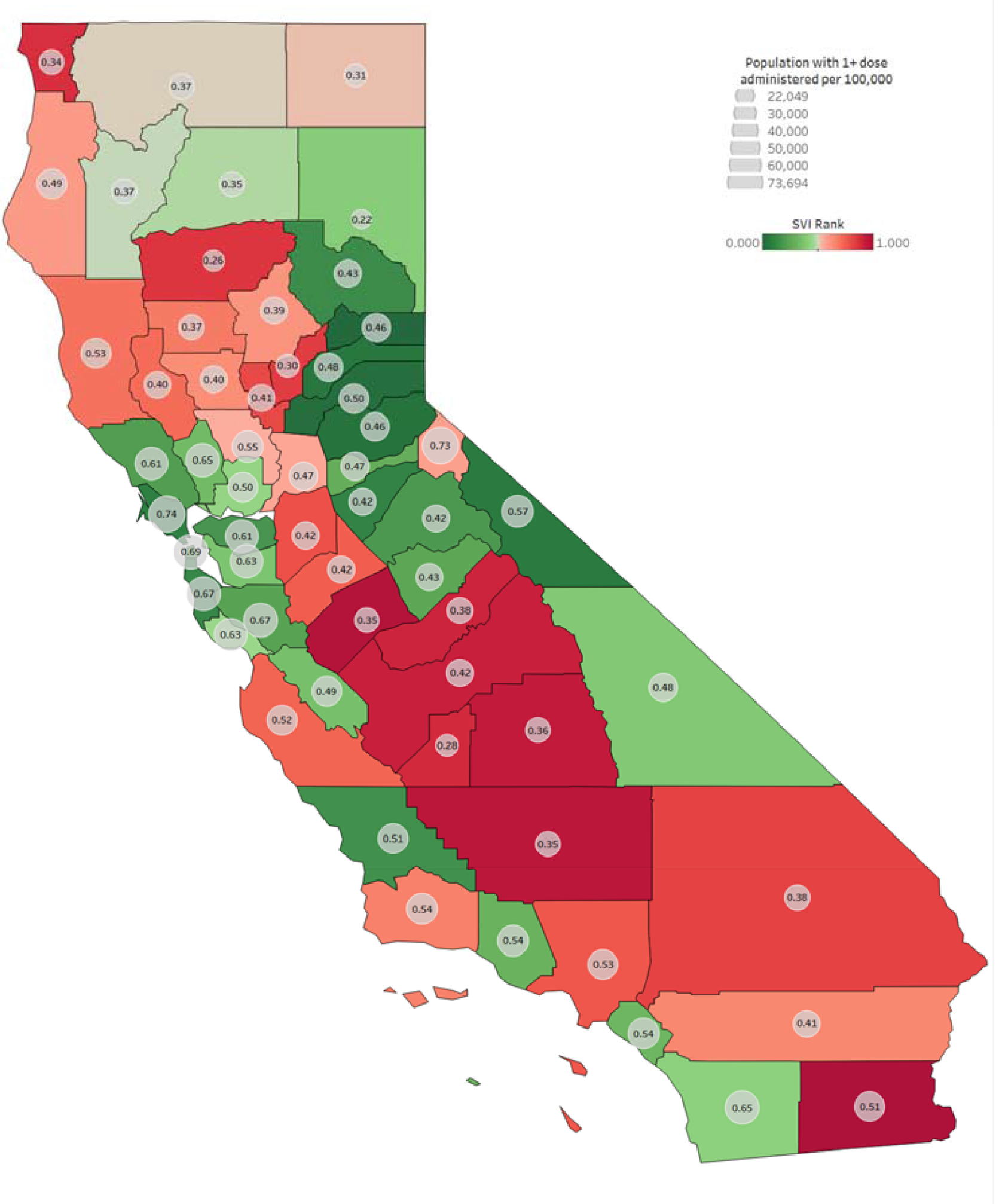
Distribution of overall Social Vulnerability Index ranks across 58 California counties and the cumulative rate of individuals who received at least one dose of vaccine per 100,000 capita (circles) at the study endpoint. Numbers in the circles represent the vaccination rate.

### Model performance

The nonlinear ecological growth curve approach allowed us to conduct meaningful hypothesis testing of the v and K parameters across counties of differing SVI status. While allowing for more flexible modeling, the use of cubic splines to describe longitudinal curves does not allow the testing of differences in growth and asymptotic maximum parameters based on SVI. Therefore, to assess the acceptability of our models, we compared their fit to the fit of alternate cubic spline models. The R^2^ values of the ecological growth curve models were compared to 1) the “gold standard” cubic spline models and 2) the “null” ecological growth curve model without an effect of SVI status. The R^2^ values of the fit ecological growth models were nearly identical to the cubic spline models, suggesting these models fit the data as well as the more complex alternative. Additionally, the R^2^ values for the models with the effect of SVI (R^2^ ranged from 0.816-0.877) were higher than the null model (R^2^=0.813), suggesting meaningful differences in longitudinal curves among counties of differing SVI status. Overall, the inclusion of SVI Theme 2 led to the best explanation of differences in proportion vaccinated across time (R^2^=0.877), while SVI Theme 4 was the poorest at explaining why proportion vaccinated differed among counties across time (R^2^=0.816). Residuals for each of the models appeared normally distributed with no outliers. We found two counties that consistently had standardized residuals greater than 3 in magnitude: Alpine County, which had consistently high proportion vaccinated versus predicted, and Lassen County, which had consistently low proportion vaccinated versus predicted. These counties did not appear to have a large effect on the estimate of the mean vaccination proportion. The final models are depicted in Figure 4.

**Figure 4.**
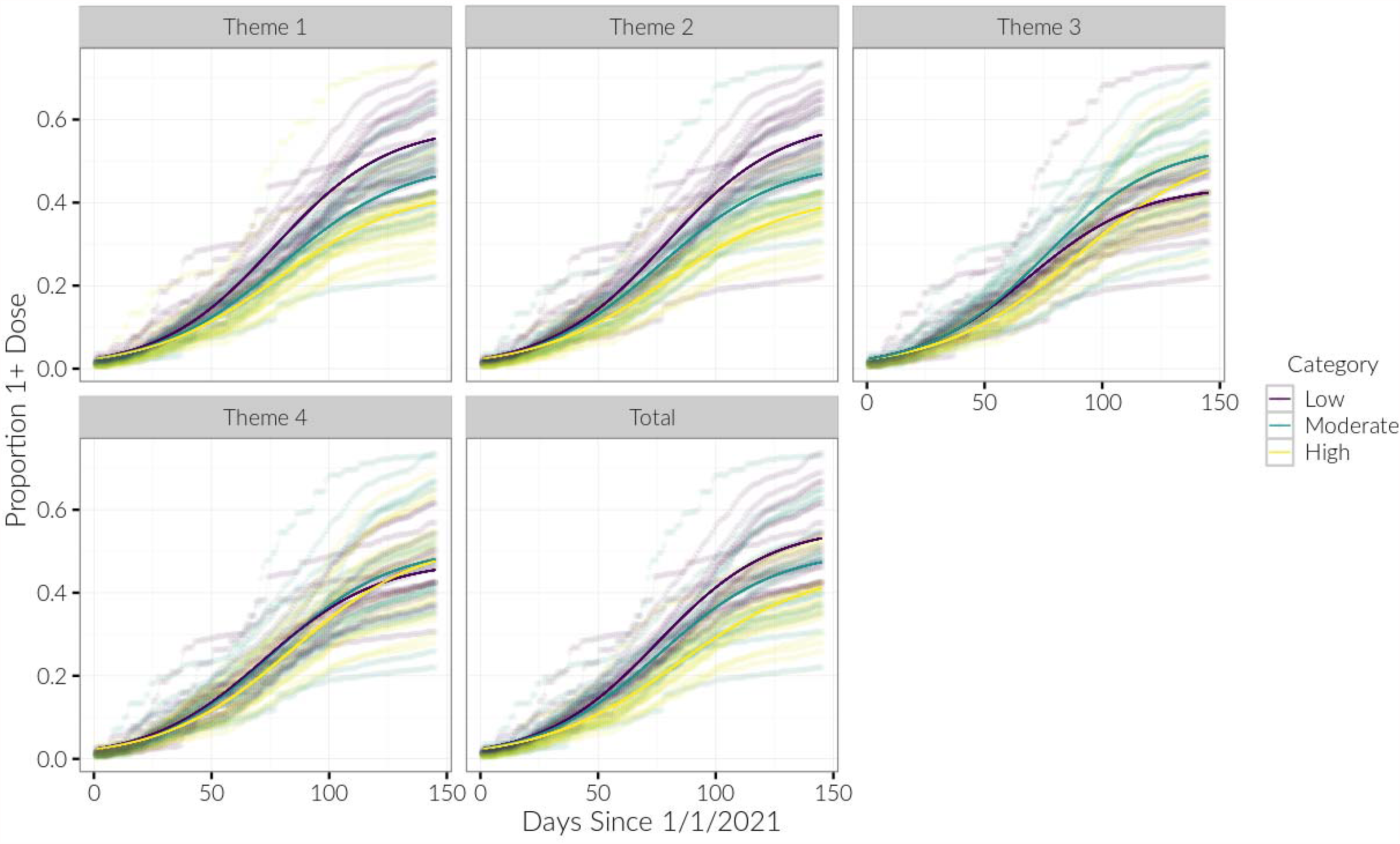
Proportion of persons with 1+ vaccination dose is shown for each county, with the model-fit estimates for each Social Vulnerability Index (SVI) category shown in bold

### Vaccination Coverage Velocity (v)

The v parameter estimates, reflecting the coverage velocity during the study period, were significantly different among the overall SVI categories, indicating an overall gap in coverage across the SVI categories over time (Figure 4, Table 1). A similar trend was observed for Themes 1, 2, and 4. However, the velocity of vaccination coverage was comparable between low and moderate categories when counties were categorized according to Theme 3 (v=0.042 and 0.041, respectively). Counties within the low category of this theme, representing those with a larger proportion of non-minority and English-speaking individuals, demonstrated the highest coverage velocity across all theme-category combinations. Meanwhile, in comparison to other themes, grouping counties by Theme 3 demonstrated the largest disparity in coverage velocity between the low and high vulnerability categories (v=0.042 vs. 0.035 respectively, *P*<0.001).

**Table 1.** Parameter estimates for each SVI category, for each theme, with 95% confidence intervals.

| Parameter / SVI Category | Overall | Theme1 | Theme2 | Theme3 | Theme4 |
| --- | --- | --- | --- | --- | --- |
| **$N_0$** |  |  |  |  |  |
|  | 0.0238 (0.0223, 0.0252) | 0.0239 (0.0225, 0.0254) | 0.024 (0.0227, 0.0254) | 0.0229 (0.0213, 0.0244) | 0.0231 (0.0215, 0.0247) |
| **$v$** |  |  |  |  |  |
| **Low** | 0.0414 (0.0403, 0.0425) | 0.0413 (0.0402, 0.0423) | 0.0404 (0.0394, 0.0414) | 0.0423 (0.0409, 0.0436) | 0.0412 (0.0398, 0.0426) |
| **Mod** | 0.0396 (0.0385, 0.0408) | 0.0376 (0.0365, 0.0387) | 0.0392 (0.0381, 0.0402) | 0.0413 (0.0401, 0.0426) | 0.0399 (0.0386, 0.0413) |
| **High** | 0.0346 (0.0334, 0.0358) | 0.0364 (0.0352, 0.0376) | 0.0355 (0.0343, 0.0366) | 0.035 (0.0338, 0.0362) | 0.0365 (0.0353, 0.0378) |
| **$K$** |  |  |  |  |  |
| **Low** | 0.561 (0.551, 0.571) | 0.587 (0.577, 0.597) | 0.602 (0.592, 0.612) | 0.441 (0.432, 0.45) | 0.478 (0.468, 0.488) |
| **Mod** | 0.504 (0.494, 0.515) | 0.502 (0.491, 0.514) | 0.501 (0.491, 0.51) | 0.542 (0.531, 0.552) | 0.512 (0.5, 0.523) |
| **High** | 0.462 (0.448, 0.476) | 0.436 (0.425, 0.447) | 0.425 (0.413, 0.436) | 0.544 (0.527, 0.561) | 0.529 (0.514, 0.544) |
| **$R^2$** | 0.857 | 0.867 | 0.877 | 0.833 | 0.816 |
| **$R^2$ Null** | 0.813 | 0.813 | 0.813 | 0.813 | 0.813 |
| **$R^2$ Spline** | 0.858 | 0.867 | 0.878 | 0.834 | 0.817 |

The $N_0$ parameter was modeled as static across the SVI categories. $R^2$ values are listed for the current models, the null model (i.e., logistic growth without an effect of SVI), and a cubic spline model. Within each model, the parameter estimate was always significantly different ( $P < .05$ ) among SVI categories except for the following: 1) in Theme 3, $v$ was not different between Low and Mod categories, 2) in Theme 3, $K$ was not different between Mod and High categories.

### Vaccination Asymptotic Maximum (K)

As demonstrated in Figure 5 and Table 1, based on the current trajectory, the asymptotic maximum for counties in the low overall SVI category is estimated to be 0.561 (95% CI: 0.551, 0.571). This parameter was lower in counties with moderate (K=0.504, 95% CI: 0.494, 0.515) and high (K=0.462, 95% CI: 0.448, 0.476) overall SVI. The models demonstrated similar patterns for themes 1 and 2. However, based on the models constructed using themes 3 and 4, there was an inverse relationship between the vulnerability categories and K estimates. Among models with differing SVI themes, the parameter estimate of K was always different (all *P*s<0.001) among vulnerability categories, except for Theme 3 which yielded comparable K values between moderate and high categories (*P*=0.06).

**Figure 5.**
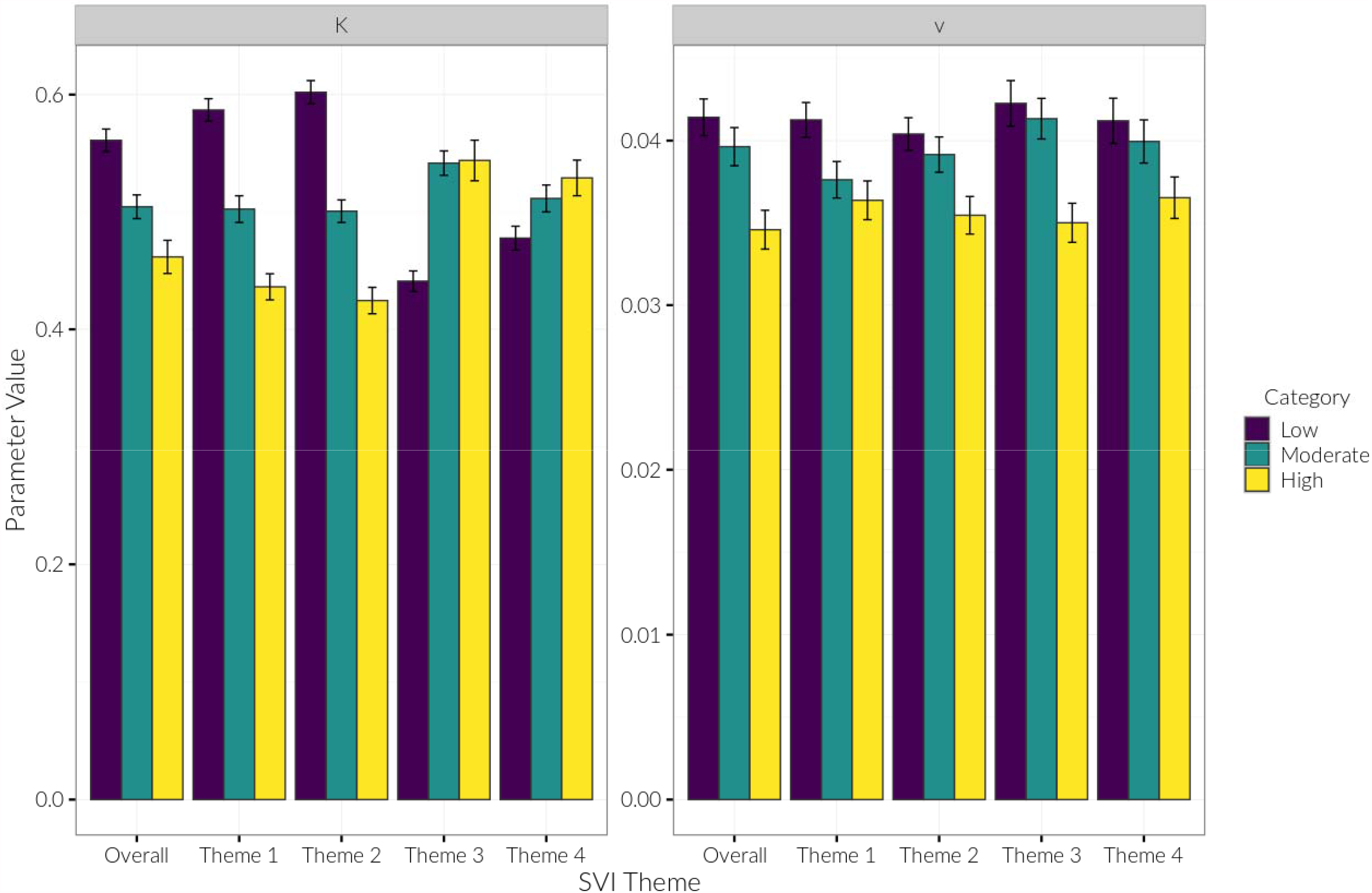
Model estimates of K and v parameters for each theme and Social Vulnerability Index (SVI) category, with 95% confidence intervals.

Among all theme-category combinations, counties in low vulnerability category of Theme 2 were estimated to achieve the largest asymptotic maximum (K=0.602, 95% CI: 0.592, 0.612), while themes 1 and 2 yielded the largest disparity: when counties were categorized according to Theme 1, the K parameters for low and high categories were 0.587 (95% CI: 0.577, 0.597) and 0.436 (95% CI: 0.425, 0.447), respectively (*P*<0.001).

### “Snapshots” Analysis

Overall, we observed large initial disparities in vaccination rates by SVI status on January 1, which appeared to become attenuated over time. For overall SVI status, on January 1, low vulnerability counties had 1.75 (95% CI: 1.44, 2.12) times the cumulative vaccination rates as high vulnerability counties (Figure 6, Table 2). This disparity significantly decreased to 1.35 (95% CI: 1.11, 1.63) times the cumulative vaccination rates by May 1 (*P*=0.03).

**Table 2.** Snapshot analysis of trends in rate ratio of proportion vaccinated in low and moderate versus high SVI counties using negative binomial regression models.

| SVI theme / Date | Vaccination Rate Ratio (95% CI) |  |
| --- | --- | --- |
|  | Low vs. High | Moderate vs. High |
| **Overall SVI** |  |  |
| January 1 | 1.75 (1.44, 2.12) | 1.56 (1.28, 1.89) |
| March 1 | 1.55 (1.28, 1.88) | 1.36 (1.12, 1.64) |
| May 1 | 1.35 (1.11, 1.63)* | 1.19 (0.99, 1.45) |
| **Theme 1** |  |  |
| January 1 | 1.91 (1.58, 2.31) | 1.37 (1.13, 1.65) |
| March 1 | 1.47 (1.22, 1.78)* | 1.17 (0.98, 1.41) |
| May 1 | 1.42 (1.18, 1.71)* | 1.16 (0.96, 1.40) |
| **Theme 2** |  |  |
| January 1 | 1.83 (1.52, 2.21) | 1.60 (1.32, 1.93) |
| March 1 | 1.46 (1.21, 1.76) | 1.26 (1.05, 1.52) |
| May 1 | 1.49 (1.23, 1.79) | 1.23 (1.02, 1.49) |
| **Theme 3** |  |  |
| January 1 | 1.42 (1.15, 1.75) | 1.62 (1.32, 1.99) |
| March 1 | 1.33 (1.08, 1.64) | 1.45 (1.18, 1.78) |
| May 1 | 0.95 (0.77, 1.17)* | 1.13 (0.92, 1.39) |
| **Theme 4** |  |  |
| January 1 | 1.16 (0.92, 1.46) | 1.03 (0.82, 1.30) |
| March 1 | 1.21 (0.96, 1.52) | 1.16 (0.92, 1.46) |
| May 1 | 0.99 (0.79, 1.25) | 1.04 (0.83, 1.30) |

\* Indicates a temporal reduction in the rate ratio for the given date versus January 1st, 2021, at $P < 0.05$ .
SVI, Social Vulnerability Index

**Figure 6.**
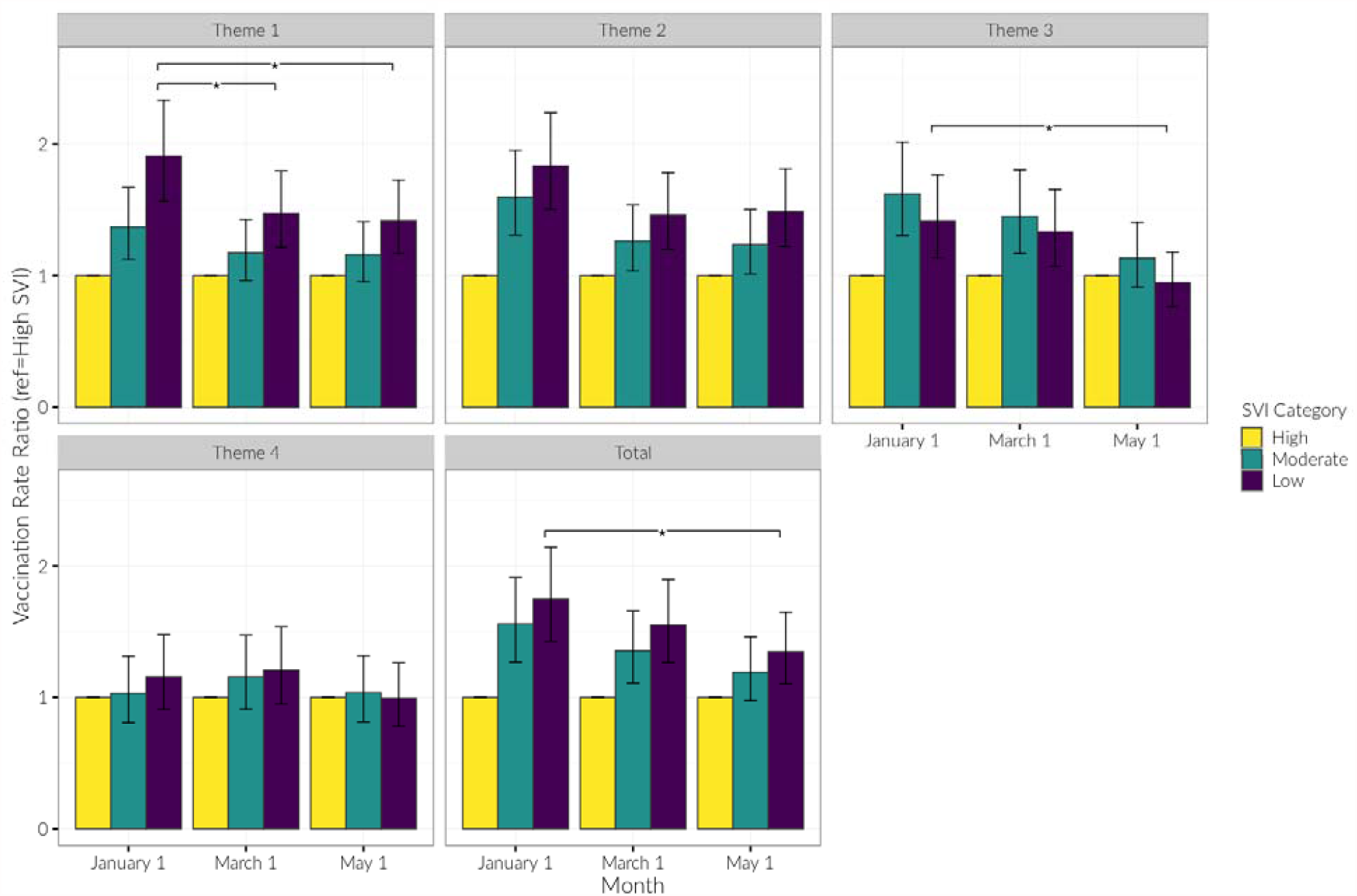
Vaccination rate ratios for low and moderate SVI vs. high SVI counties, by date and SVI category. Presented are rate ratios with 95% confidence intervals. * indicates a temporal reduction in the rate ratio for the given date vs. January 1 at p < 0.05.

There was additionally a large initial disparity in cumulative vaccination rates by SVI Theme 1. On January 1, low vulnerability counties had 1.91 (95% CI: 1.58, 2.31) times the cumulative vaccination rates as high vulnerability counties, which decreased to 1.42 (95% CI: 1.18, 1.71) times the rate by May 1 (P=0.01). With regard to SVI Theme 3, low vulnerability counties began the year with 1.42 (95% CI: 1.15, 1.75) times the cumulative vaccination rates as high vulnerability counties, but by May 1 this rate ratio had decreased (*P*=0.002) and there was effectively no difference in cumulative vaccination rates by SVI Theme 3 (RR=0.95, 95% CI: 0.77, 1.17). Trends in decreasing rate ratios over time, suggesting more equal cumulative vaccination rates by SVI status over time, were seen for the other themes.

## 4. Discussion

The COVID-19 pandemic shed a broader light on the existing gaps in healthcare delivery and outcomes across the socio-demographic subgroups in the U.S. This study was performed to determine the vaccination coverage velocity of California counties based on the underlying social vulnerability status, and to utilize the longitudinal trends in vaccination to estimate the disparities in maximum anticipated proportion of vaccinated individuals based on the current trajectory. Using the data from 58 California counties, we demonstrated a decreasing yet significant gap in vaccination coverage based on the overall SVI status. Counties with high vulnerability designation in terms of minority status and language (Theme 3) demonstrated the lowest coverage velocity across all theme-category combinations. Meanwhile, counties in this category have suffered from the highest average mortality rate as of May 23, 2021, when compared to moderate and low categories.

The large initial disparities in vaccination rates by overall SVI status, Theme 1, and Theme 3 attenuated over time, while counties in the high vulnerability category according to themes 3 and 4 are estimated to eventually achieve a higher proportion of residents vaccinated compared to less vulnerable counterparts. Given the higher proportion of Black and Hispanic populations in the high vulnerability category of Theme 3, our findings about this theme parallel those of a study of 3138 U.S. counties which showed that counties with large Hispanic and Black populations have a higher vaccination uptake^13^. It seems that when these populations faced challenges to accessibility, a low vaccine coverage velocity was observed, while improved accessibility led to a greater uptake.

The higher estimated asymptotic maximums in vulnerable counties can possibly be attributed to California-funded target outreach programs, media spending, and increased vaccine allocation targeted toward the most vulnerable populations often consisting of higher minority populations. As demonstrated in Figure 4, close to 100 days after January 1, 2021, the increase in proportion of residents vaccinated in low and moderately vulnerable counties according to Theme 3 began to show a plateau, while the pace of coverage was relatively maintained in highly vulnerable counties. Additionally, the vaccination rate ratios for low and moderate SVI Theme 3 counties diminished from March to May, reaching close to 1.0 in reference to high vulnerability counties. This change in trend occurred nearly concomitantly with California’s fervent spending in community outreach programs ($30 million), media ($40 million), and increased vaccine allocation for the hardest-hit communities^14^ in March 2021. Similar findings have been reported in Maryland, where the state’s Vaccine Equity Task Force used a targeted approach to improve the Vaccine Equity Index (VEI) of Black adults aged 65 and older by 48% and Hispanic/Latinx adults aged 65 and older by 36% between February 17, 2021, and April 7, 2021^15^, which suggests that targeting communities based on vulnerability metrics can help mitigate health inequities.

In addition to suffering a myriad of consequences brought upon by COVID-19, residents of counties classified as high vulnerability within Theme 3 (minority status and language) share overlapping characteristics with residents of counties classified as high vulnerability within Theme 4 (housing type and transportation). Racial/ethnic minority populations are more likely to live in multigenerational households^16^ or in more crowded neighborhoods and units^17^. Latinx persons in California are 8.1 times more likely to live in households facing exposure risks like living in households with essential workers and having fewer rooms than household members^18^. These overlapping characteristics may explain our observed similarity in trends of asymptotic maximum between themes 3 and 4.

Vaccine disparities across all SVI themes have decreased over time (Figure 6) likely due to accessibility becoming more ubiquitous with programs targeting vulnerable populations. The early disparities in vaccination rates observed for themes 1 and 3 reflect the fact that neither socioeconomic nor racial/ethnic minority status was initially the basis for vaccine prioritization. Due to the limited supply, the COVID-19 vaccines were distributed across the U.S. in phases, prioritizing healthcare workers, long-term care residents and staff as well as persons aged 65 and older. One possible explanation for this significant initial disparity lies in the racial composition of the U.S. adults aged 65 and older, with 76% being White while only 10% being Black in 2019 according to the U.S. Census^19,20^. Other factors such as complex registration protocols, technological illiteracy, and transportation barriers likely created an unnecessary delay in the vaccination process^21^, particularly for residents of high vulnerability counties pertaining to Theme 1 and Theme 3, who are more likely to lack resources that facilitate the process of getting a vaccine^22^.

To mitigate the vaccine inequities, elimination of barriers to vaccination is just as important, if not more, than setting equitable vaccine eligibility and prioritization criteria. Individuals of an ethnic/racial minority are typically over-represented among those with essential public-facing occupations including the services and transportation^17^. Such occupations were prioritized during the initial vaccination phases, yet many workers were likely unable to receive the vaccine due to a multitude of barriers, such as concerns about time off of work. Furthermore, uninsured individuals may have had concerns about the healthcare costs associated with vaccine side effects, while immigrants may have faced unique challenges due to the vaccination data being tracked by personal identifiers, fearing their immigration status would be jeopardized^23^.

Although many studies suggest inaccessibility as a significant factor behind vaccination inequities, the negative contribution of vaccine hesitancy cannot be fully discounted, especially among minorities^15^. A longitudinal survey of COVID-19 vaccine acceptance showed that in November 2020, only 42% of African American respondents expressed their willingness to receive the vaccine, compared to 61% of White respondents^24^. Another study conducted between March 24, 2020, and February 16, 2021, yielded similar results; compared with White participants, the odds ratios for vaccine hesitancy were 3.15 for Black participants and 1.42 for Hispanics^25^. However, although earlier vaccine hesitancy surveys can potentially explain the initial higher vaccine disparities among the Theme 3 categories, more recent surveys show similar levels of willingness to get vaccinated across Black, Hispanic, and White populations^26–28^, which could be a contributing factor to the amelioration of vaccine inequity witnessed in high vulnerability Theme 3 counties from March to May 2021 (Figures 4 and 6). These increased levels of willingness to get vaccinated among racial/ethnic minority groups can potentially be a result of outreach programs like lotteries and mobile vaccine clinics^29^. While a recent study has shown that misinformation can lead to significant declines in willingness to get vaccinated^30^, California’s outreach and media spending consist of public education programs in efforts to combat such misinformation and better educate more vulnerable residents, which can also lead to increases of willingness to get vaccinated^14^.

Our study showed that the SVI-overall and its four themes are useful measures in distinguishing differing vaccination rates across counties. As SVI identifies counties with the most vulnerable populations, it can be used to help mitigate health inequities as recommended by The National Academies of Sciences, Engineering, and Medicine (NASEM)^31^. This measure has been adopted by 29 jurisdictions (28 states and one city) across the United States to guide equitable vaccine distribution during the COVID-19 pandemic^32,33^. Based on our findings, our model for Theme 2 (household composition and disability) demonstrated the best performance, while Theme 3 (minority status and language) yielded the highest discriminative value in terms of vaccination coverage velocity, suggesting this theme as the most informative underlying domain of SVI in the context of COVID-19 vaccination. While the generalizability of this finding is unknown, more empirical data are needed to unfold the contribution of each SVI category to health outcomes.

## 5. Limitations

Despite our best efforts, this study is not without limitations. In the early phase of the COVID-19 vaccine rollout, only specific populations, such as healthcare/frontline workers and people with underlying health conditions were eligible for vaccination. This staged vaccine rollout may have affected the study findings, particularly those related to SVI Theme 2, since counties with a higher proportion of individuals older than 65 may have achieved a higher early vaccination coverage. Moreover, the SVI does not include some of the underlying factors that potentially contributed to disproportionate vaccination coverage. For instance, internet access and digital literacy are vital for scheduling vaccine appointments and can facilitate this process. Individuals older than 65, low-income individuals, and Black and Hispanic individuals are more likely to lack internet access through computers^22^, which further exacerbates vaccine inequity issues. In terms of dataset limitations, from December 15, 2020, to May 23, 2021, daily updated vaccine numbers were missing in some spatio-temporal data points and were handled by imputing the latest available day’s data for the respective county. However, given the relatively small number of missing data points and the nature of our analysis, this limitation is unlikely to have introduced significant bias. As new data become available, future analyses should confirm that vaccination trajectories actually approach the estimated asymptotic maximums predicted in this study.

## 6. Conclusion

The present study provides insight into the problem of COVID-19 vaccine disparity across California which can be used to help promote equity during the current pandemic and guide the allocation of future vaccines such as COVID-19 booster shots. By addressing the relationship between SVI (and its four components) with longitudinal trends of vaccine administration, we demonstrated its utility in the development of target-based equitable strategies.

## Data Availability

Publicly available data was used in the process of this study.

## 7. Declarations

### Ethics approval

Ethical approval was not acquired nor applicable for this research article. No human or animal participants were involved in the study. Publicly available data was used in the process of this study.

### Data availability

The data underlying this article will be shared on reasonable request to the corresponding author.

### Funding

This study was supported by the National Science Foundation (NSF) under Award Number 2027456 (COVID-ARC). This publication was supported by grants UL1TR001855 and UL1TR000130 from the National Center for Advancing Translational Science (NCATS) of the U.S. National Institutes of Health (NIH). The content is solely the responsibility of the authors and does not necessarily represent the official views of the NSF, NIH, or other entities.

### Conflict of Interest

None declared.

## Notes

### Competing Interest Statement

The authors have declared no competing interest.

### Author Declarations

Ethical approval/IRB was not acquired nor applicable for this research article. No human or animal participants were involved in the study. Publicly available data was used in the process of this study.

